# Whole genome sequencing of hepatitis B virus (HBV) using tiled amplicon (HEP-TILE) and probe-based enrichment on Illumina and Nanopore platforms

**DOI:** 10.1101/2024.09.11.24313306

**Authors:** Sheila F Lumley, Chris Kent, Daisy Jennings, Haiting Chai, George Airey, Elizabeth Waddilove, Marion Delphin, Amy Trebes, Anna L McNaughton, Khadija Said Mohammed, Sam Wilkinson, Yanxia Wu, George MacIntyre-Cockett, Beatrice Kimono, Moses Kwizera, Kevin Ojambo, Tongai G Maponga, Catherine de Lara, Jacqueline Martin, James Campbell, Marije Van Schalkwyk, Dominique Goedhals, Robert Newton, Eleanor Barnes, Nicholas J Loman, Paolo Piazza, Joshua Quick, M Azim Ansari, Philippa C Matthews

## Abstract

Hepatitis B virus (HBV) whole genome sequencing (WGS) is currently limited as the DNA viral loads (VL) of many clinical samples are below the threshold required to generate full genomes using current sequencing methods. We developed two pan-genotypic viral enrichment methods, using probe-based capture and tiled amplicon PCR (HEP-TILE) for HBV WGS. We demonstrate using mock samples that both enrichment methods are pan-genotypic (genotypes A-J). Using clinical samples, we demonstrate that HEP-TILE amplification successfully amplifies full genomes at the lowest HBV VL tested (30 IU/ml), and the PCR products can be sequenced using both Nanopore and Illumina platforms. Probe-based capture with Illumina sequencing required VL >300,000 IU/ml to generate full length HBV genomes. The capture-Illumina and HEP-TILE-Nanopore pipelines had consensus sequencing accuracy of 100% in mock samples with known DNA sequences. Together, these protocols will facilitate the generation of HBV sequence data, enabling a more accurate and representative picture of HBV molecular epidemiology, cast light on persistence and pathogenesis, and enhance understanding of the outcomes of infection and its treatment.

## Manuscript

### 1. Introduction

Worldwide an estimated 254 million people are living with chronic HBV infection (CHB) [1]. Despite the availability of prophylactic vaccines and suppressive antiviral therapies, the annual toll of approximately 1.1 million HBV-related deaths in 2022 [1] highlights the persistent global challenge faced. HBV whole genome sequencing (WGS) provides genetic insights into the pathogen at population and individual levels [2]. At a population level, WGS improves our understanding and monitoring of HBV drug resistance, vaccine efficacy, diagnostic escape and tracking of outbreaks. In individuals, WGS has the potential to contribute to personalised healthcare, e.g. informing modification of antiviral regimens to take into account antiviral resistance associated mutations (RAMs), and risk-stratified hepatocellular carcinoma (HCC) surveillance [3].

There are several challenges associated with HBV WGS. Target enrichment using probe based capture, HBV specific polymerase chain reaction (PCR) or rolling circle amplification (RCA) [4,5] is required when performing HBV WGS due to frequently low DNA viral loads (VL) [6], the tiny genome (3.2kB), and abundant host nucleic acid background. Sequencing low VL samples, although challenging, is important at both an individual and population level, in order to understand mechanisms of transmission, persistence, immunological control and clearance, breakthrough viraemia on treatment, and for characterising occult HBV infection. HBV has ten distinct genotypes (A–J), with >8% nucleotide divergence, which are geographically distributed and associated with infection outcomes [7]. This diversity leads to challenges in designing pan-genotypic capture probes and PCR primers. The structure of the HBV genome also presents challenges for WGS. In the blood, HBV DNA is present predominantly in a relaxed-circular (rc-DNA) form, a circular, partially double-stranded (ds)DNA configuration (**Figure 1A**) [8]; neither positive nor negative DNA strand is continuous.

**Figure 1:**
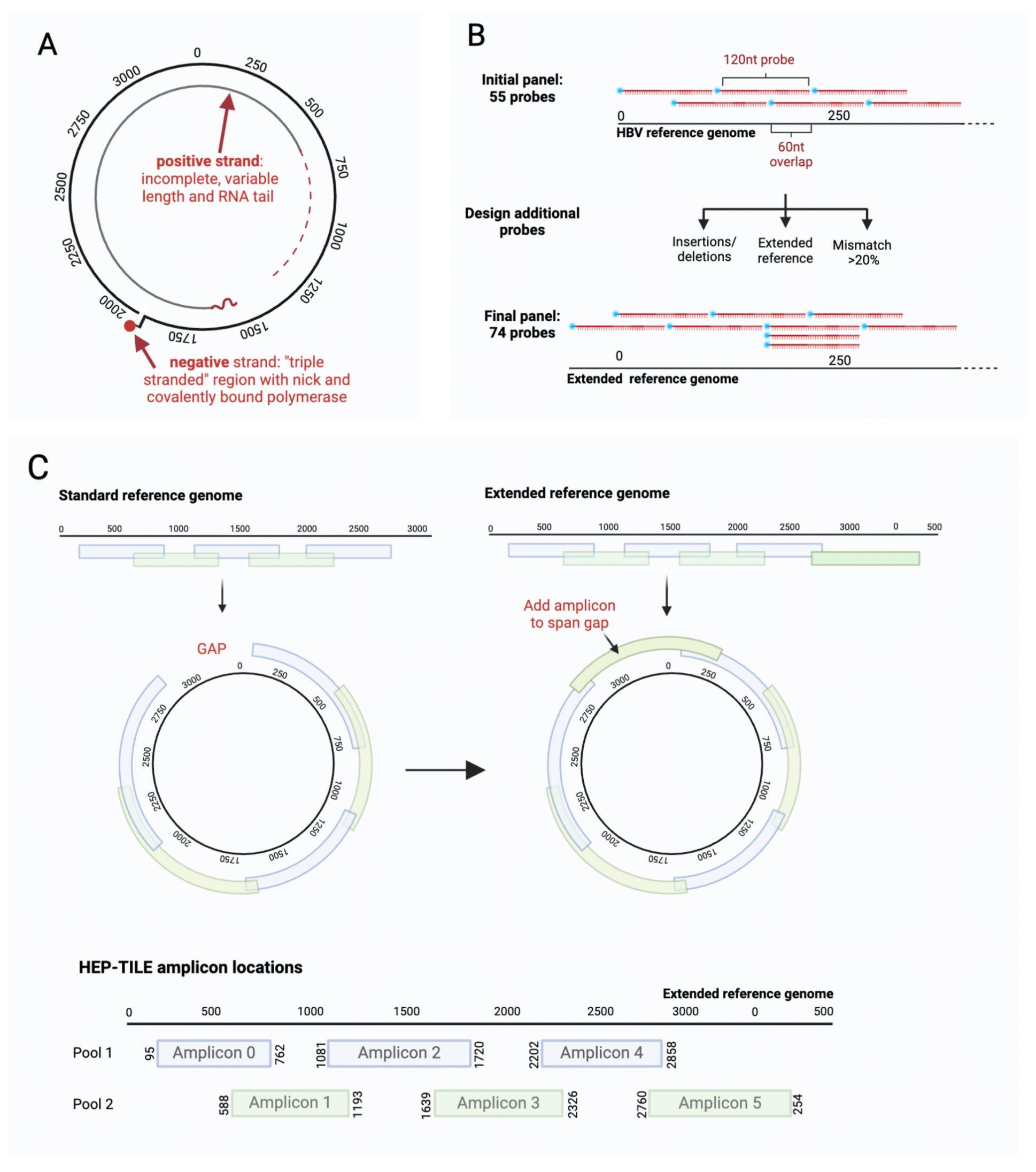
Design of HBV targeted enrichment methods. (A) HBV genome structure: relaxed circular DNA (rc-DNA). The complete negative strand is discontinuous, with a ‘nick’ and covalently bound polymerase. The positive strand is incomplete with a variable length and RNA tail. (B) Probe design schematic for HBV probe based enrichment. An initial set of probes are designed based on non-recombinant whole genomes downloaded from HBVdb. Additional probes are added in regions of high diversity, using an extended reference sequence to account for the circular genome, and to account for insertions/deletions, then the final probe panel confirmed. (C) Multiplex tiling PCR for the circular HBV genome. Generation of tiled schemes from linear reference genomes leaves a gap in coverage for a circular genome. An additional amplicon spanning the start/end of the linear reference ensures a theoretical maximum 100% genome coverage can be obtained. Primer positions and amplicon length for HBV tiled amplicon scheme. Numbering of nt positions based on X02763 reference genome. Created in BioRender. Lumley, S. (2024) BioRender.com/r40b439

We set out to optimise and validate workflows for HBV WGS. Due to limitations in sensitivity of our RCA protocol [4] (related to difficulty in generating a fully circular template), we initially used probe-based enrichment, a strategy which has previously been validated across diverse pathogens [9–11]. The method is based on the design of biotinylated single-stranded DNA probes, designed to hybridise with target viral sequences within a sequencing library, and selectively capture HBV DNA, thereby enriching the final library for HBV DNA and increasing on-target sequencing yield (**Figure 1B**).

Second, due to limitations in sensitivity of probe-based enrichment, we developed a tiled amplicon approach for HBV whole genome sequencing (HEP-TILE) (**Figure 1C**) using HBV-specific primers designed using PrimalScheme3, successor to PrimalScheme [12,13]. PrimalScheme3 includes key updates to allow the generation of WGS from pathogens with circular genomes, with primers able to deal with greater pathogen diversity. The HBV sequencing wet and dry lab workflows build on the widely used ARTIC-Network nCoV-2019 sequencing protocol [14] and fieldbioinformatics [15], both of which have been used extensively worldwide during the SARS-CoV-2 pandemic.

Scale up of WGS requires development of open access, easily deployable sequencing methods ideally building on existing infrastructure and expertise. By developing the protocols described here, we aim to facilitate the generation of HBV sequence data globally, to obtain a more accurate and representative picture of HBV genetics and molecular epidemiology.

## 2. Methods

### 2.1 Samples

#### 2.1.1 Mock samples

We prepared mock HBV samples using plasmids containing ∼1.3 copies of the HBV genome representing genotypes A-J in a pUC57 backbone, (genotypes A-F from [16], G-J designed in-house and produced by GeneArt) (**Supplementary table 1**). We spiked plasmids into non-infected human serum (Merck, H5667) at a concentration of 10^5^ plasmid copies/ml.

#### 2.1.2 Clinical samples

We used 63 plasma samples collected between 2011-2023 from adults with CHB in three settings: (1) outpatient clinics in Oxford (UK) [6,17], (2) outpatient clinics in Tygerberg Hospital, Cape Town (Stellenbosch University) and public sector hospital services in Bloemfontein (University of the Free State), South Africa [6], and (3) participants of the Uganda Liver Disease Study (Kalungu District, south west Uganda [18,19]). We selected samples to represent a range of VL and genotypes; methods were developed iteratively and based on available samples. (**Figure 2, Supplementary table 2**).

**Figure 2:**
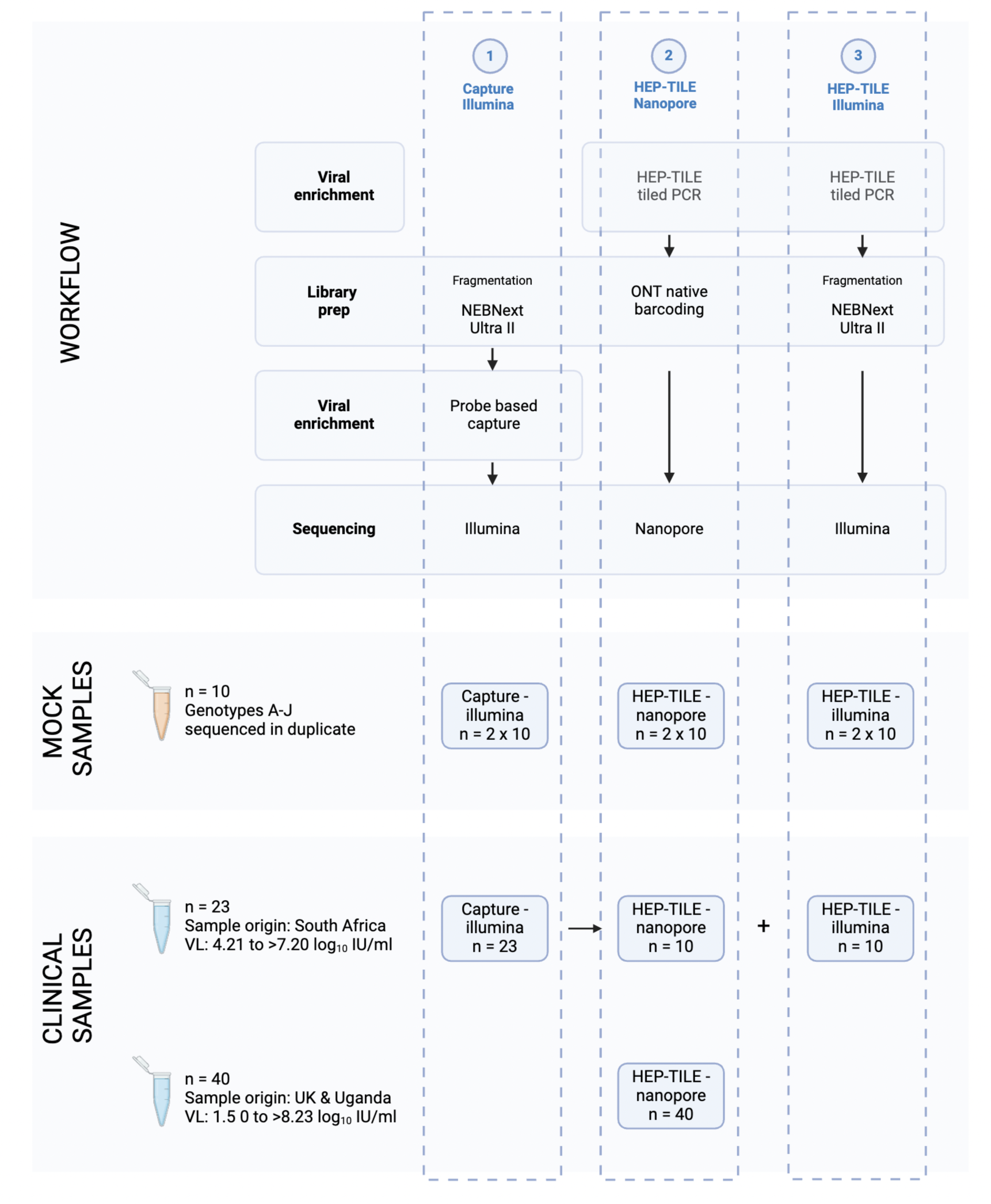
Overview of sequencing workflows. Numbers of samples sequenced at each stage indicated. Due to sample availability, project stage, and local infrastructure, we used different subsets of samples to validate each method. In the clinical sample set from South Africa, if residual extract was available after capture, two further aliquots were taken forward for HEP-TILE enrichment and sequencing on Nanopore and Illumina. Created in BioRender. Lumley, S. (2024) BioRender.com/k41p399

Written informed consent was provided by participants at enrollment. Approval for this work was provided as follows, all methods were performed in accordance with the relevant protocols:

● UK patients: Oxford Research Ethics Committee A (ref. 09/H0604/20)
● South Africa / Stellenbosch patients: Stellenbosch University (ref. N17/01/013) and OxTREC (ref 1-18)
● South Africa / Bloemfontein patients: University of the Free State (ref. UFS-HSD2018/0193-0001) and OxTREC (ref 1-18)
● Uganda patients: Uganda Virus Research Institute Research and Ethics Committee (ref. GC/127/19/04/711), Uganda National Council for Science and Technology, and Oxford Tropical Research Ethics Committee (ref 50-18)

We collected blood samples in EDTA tubes. To separate plasma, we centrifuged whole blood at 1800 for 10 minutes. We harvested the supernatant and stored it at –80°C.

Quantification of HBV DNA VL was performed on the clinically validated Abbott M2000 platform by Oxford University Hospitals (OUH) diagnostic microbiology laboratory for UK samples, and with the Cobas Ampliprep/Taqman HBV test for Ugandan and South African samples (performed by by the Clinical Diagnostic Laboratories for MRC/UVRI/LSHTM Research Institute Uganda and the NHLS Tygerberg Virology Laboratory respectively). VL ranged from 1.5 to > 8.3 log_10_ IU/ml (i.e. above the assay limit of quantification) (**Supplementary table 2**).

#### 2.1.3 Host depletion and nucleic acid extraction

For mock and clinical samples initially being enriched by capture, we performed host depletion with micrococcal nuclease (NEB catalogue number M0247S, 250µl plasma with 1.25µl MNase and 23.75µl MNase buffer incubated for 20 mins at 37°C) followed by extraction using the Kingfisher Apex Magmax viral/pathogen nucleic acid isolation kit (two aliquots of MNase-treated plasma eluted into 50µl of kit elution buffer, combined followed by a SPRIselect bead clean up to reduce volume to 40µl).

For samples from UK and Uganda for HEP-TILE Nanopore sequencing, we extracted DNA using the QIAgen MinElute Virus Spin kit from either 200µL or 400µL input plasma. Double volume (400µl) extraction was used if VL was <3 log_10_ IU/ml (with double volume protease, ethanol and buffer AL). We used carrier RNA as per manufacturer’s instructions, and eluted into 45µl H_2_O. The sample volume and extraction method used for each sample is detailed in **Supplementary table 2**.

### 2.2 Public data for probe/primer design

We based our probe and primer design on downloads of all available HBV non-recombinant whole genomes from the Hepatitis B Virus Database (HBVdb) [20] on 31st January 2019 for probe design and 15th February 2024 for primer design (**Supplementary table 3**). In addition for HEP-TILE, genomes for genotypes I and J were sourced from McNaughton et. al [7].

### 2.3 HBV probe based enrichment

#### 2.3.1. Probe design

We used RaxML [21] with a general time reversible model with Gamma model of rate heterogeneity (“-m GTRGAMMA” option in RAxML) to infer a maximum likelihood phylogeny of HBV full genomes. Next, we used RAxML to infer the ancestral sequences and as input we used the midpoint rooted tree and the sequence of our isolates with the GTRGAMMA option. We used the ancestral sequence at the root of the tree to design the first set of probes assuming that this sequence on average has the least amount of divergence relative to all other isolates.

As the HBV genome is circular, we added 120 bases from the beginning of the ancestral root sequence to the end of the sequence to ensure that capture probes cover the break point which is used to present the genome linearly. We then divided the ancestral root sequence into 120 nucleotide (nt) segments with 60 bases overlap which resulted in an initial set of 55 probes (**Figure 1B**). Genotype G has an insertion of 36 bases relative to other genotypes in the core gene. As the ancestral root sequence contained this insertion, we also designed a probe of 120 nt which lacked this insertion. Furthermore, genotype D has a deletion of 33 bases in the pre-S1 region relative to all other genotypes. To ensure a probe covers this region for genotype D, we designed a probe of 120 nt that lacked this region.

Our previous work in HCV probe-based sequence capture demonstrated that probes of 120 bases long can tolerate up to 20% divergence relative to their target sequence before the efficiency of capture drops [11]. To ensure that the designed probes are within a maximum of 20% divergence of each viral sequence, we divided the dataset based on genotype and created a consensus sequence for each genotype. We then aligned the probes to each viral sequence and measured the proportion of mismatches between each probe and the isolate. For each viral sequence, if a continuous region of ≥60 bases diverged from the probe sequences by >20%, a new probe was designed for the region using the genotype consensus sequence. As a quality control step, we removed any potential sequences that contained an “N” as we assumed that the sequence may be of low quality. Additionally we counted the number of ambiguous nucleotides and any viral sequence containing ≥5 ambiguous nucleotides was also removed. The final probe set contained 74 probes.

The probe sequences and the set of HBV genome sequences that were used for their design are presented in **supplementary table 4** and can also be downloaded from the following webpage: https://figshare.com/articles/dataset/HBV_probe_sequences/22127015

#### 2.3.2. Library preparation, hybrid capture and sequencing protocol

We prepared libraries for Illumina sequencing using the NEBNext Ultra II FS DNA library preparation kit (protocol for use with inputs <100ng). We used an input of 26µL extracted DNA for the fragmentation/end preparation reaction and fragmented for 3 minutes, with a negative (mastermix only) control. We performed indexing PCR using NEBNext multiplex Oligos UDI and 13 amplification cycles. We pooled the amplified libraries corresponding to each aliquot in equivolume proportions to generate a final multiplex library. We purified the pool using SPRIselect beads, eluted into a final volume of 20µL and subsequently quantified using High Sensitivity dsDNA Qubit assay (Invitrogen) and Agilent 2100 Bioanalyzer high sensitivity DNA protocol.

We concentrated a 4.4µg aliquot of the final multiplexed library using the manufacturer’s AMPure XP Bead DNA concentration protocol, then enriched for HBV using the custom-designed probe panel (IDT Technologies) and xGen Hybridization and Wash Kit (IDT Technologies) following manufacturer’s ‘tube protocol’. We amplified the final enriched library (12 cycles on-bead PCR), repurified (eluting into 12µL EB), quantified using High Sensitivity dsDNA Qubit assay (Invitrogen) and Agilent 2100 Bioanalyzer high sensitivity DNA protocol, then sequenced on the Miseq v3 with 2×300nt paired-end reads or on a partial lane of Novaseq X 2×150nt paired-end reads.

#### 2.3.3. Data analysis - capture Illumina pipeline

We trimmed de-multiplexed sequence read-pairs of low-quality bases using QUASR (version 7.01) [22], trimmed adapter sequences with CutAdapt (version 4.8) [23] and Skewer (version 0.2.2) [24] and subsequently discarded if either read had <50bp sequence remaining. We mapped the cleaned read pairs to human reference genome hg19 using Bowtie2 (version 2.2.4) [25] and excluded from further analyses. All nonhuman read pairs were mapped using BWA (version 0.7.10) [26,27] to a set of 44 HBV references covering all known HBV genotypes and subgenotypes to choose an appropriate reference [7]. We chose the HBV reference with the greatest number of HBV reads mapping to it as the genetically closest reference to the sequenced isolate. Next, we re-mapped all non-human read pairs to the closest HBV reference. We then used Picard markduplicates tool (version 1.111) [28] to remove duplicate read pairs (where read pairs starting in the same place and ending in the same place on the genome are assumed to be PCR duplicates). A base was called at the consensus level if allele depth was above x5, with HBV genome coverage calculated by determining the percentage of bases called at the consensus level.

### 2.4 HBV tiled amplicon scheme (HEP-TILE)

#### 2.4.1 PrimalScheme 3

We developed a pan-genotypic HBV scheme using an early version of PrimalScheme3 (https://github.com/ChrisgKent/primalscheme3), the successor to PrimalScheme [13], a web-based primer design tool for developing multiplex primer schemes. A number of changes were made in PrimalScheme3 to enable us to generate an overlapping (tiled) amplicon scheme which covered the circular HBV genome, utilising a number of discrete primers at each position to handle intraspecies diversity.

Usually an amplicon scheme results in a short sequence at each end of the reference genome which is not covered due to primer placement constraints and downstream primer-trimming. With the conventionally designated ‘start site’ for the circular HBV genome being within the overlapping surface and reverse transcriptase genes (**Figure 1**), this results in the loss of epidemiologically and clinically relevant information. PrimalScheme3 introduces an option to add an amplicon spanning the start/end of circular genomes, resulting in 100% genome coverage.

Intra-species diversity has posed an issue for amplicon sequencing, as variation within the primer binding sites reduces primer binding efficacy. The original PrimalScheme identified conserved primer binding sites by heavily penalising those with variation within the input genomes. This approach is effective for closely related genomes, such as outbreak strains, but for diverse inputs a different approach is needed. PrimalScheme3 handles diversity with the use of ‘primer clouds’ i.e. a discrete set of 3’-anchored primers which cover all mutations above a user-specified frequency, without the use of ambiguous bases. This approach reduces the negative effects of adding additional primers to a multiplexed PCR, such as primer-primer interactions, and mispriming.

#### 2.4.2 Multiplex primer pool design for HBV (HEP-TILE)

We aligned HBVdb HBV genomes from each genotype (A-H) separately using MAFFT [29], and phylogenetically downsampled each genotype to 0.95 relative tree length using TreeMMer [30]. We combined each downsampled genptype’s genomes with genotype I and J sequences from[7], aligned with MAFFT [29], and input into PrimalScheme3 command line tool, with the options; -- ampliconsize 600, --minbasefreq 0.02, --backtrack and –minoverlap 20.

Initially, we developed a 500bp, eight amplicon scheme (https://labs.primalscheme.com/detail/hbv/500/v1.1.0) [31], however when trialled, one amplicon spanning 1700-2000 nt consistently dropped out in samples with VL <5 log_10_ IU/ml (**Supplementary figure 1A and 1B)**. We hypothesise that is explained by the structure of HBV DNA: in low VL samples only rc-DNA is present, which has discontinuities in both the positive and negative DNA strands in that region leading to amplicon dropout. In contrast, in higher VL samples, covalently closed circular DNA (cccDNA) is also present in the plasma [32,33]. cccDNA has continuous positive and negative strands which act as a template for this amplicon. We identified a suitable location for primers spanning this region, taking into account the structure of rc-DNA, and re-designed a 600bp primer scheme (hbv/600/v2.1.0) (**Supplementary figure 1C-E**), resulting in a six amplicon scheme with amplicons ∼600-715bp long, generating a theoretical 100% HBV genome coverage (**Figure 1C**). This scheme uses 131 primers to cover the HBV diversity present in our reference dataset.

#### 2.4.3 HEP-TILE PCR and sequencing protocol

Amplification and library preparation were performed by adapting the nCoV-2019 LoCost v3 sequencing protocol [14], with the removal of the reverse transcriptase step (full protocol on protocols.io [34]). Extract from samples with VL >6 log_10_ IU/ml were diluted 10 fold prior to PCR. Negative (mastermix only) controls were added at the PCR and library prep stages. We used the native barcoding kit SQK-NBD114.96 and R10.4.1 MinION flowcells (Oxford Nanopore Technologies, ONT), multiplexing up to 96 samples per run and sequencing for 72 hours. For Illumina sequencing of tiled amplicons, we fragmented the amplicons and performed library preparation using the NEBNext Ultra II FS DNA kit, multiplexing up to 96 samples per run and sequenced on a partial lane of Novaseq X 150PE.

#### 2.4.4 Data analysis - HEP-TILE Nanopore pipeline

We basecalled reads and demultiplexed data using dorado (0.7). We generated a novel pipeline hbv-fieldbioinformatics [35] to produce consensus genomes. Reads are mapped against representative genomes from all genotypes [7], with the genome with the most mapped reads being selected as the primary reference. To handle the circular genome, an intermediate circular primary reference genome is created, where the sequence of the circular amplicon is appended to the 3’ end of the reference. Reads are then remapped to the circular reference with minimap2 (2.26), primers are trimmed, followed by variant calling by LongShot and Medaka (0.4.5, 1.11.3). Variants and read depth for the appended region are then mapped back to their corresponding position on the original reference. A base was called at the consensus level if allele depth was over x20, with HBV genome coverage calculated by determining the percentage of called consensus bases.

#### 2.4.5 Data analysis - HEP-TILE Illumina pipeline

Similar to the capture-based Illumina pipeline, we initially used QUASR (version 7.01) [22] to filter out poor de-multiplexed sequencing reads. We then trimmed primer sequences by using CutAdapt (version 4.8) [23] and Skewer (version 0.2.2). Human-like reads were excluded with Bowtie2 (version 2.2.4) [25]. We extended all 44 HBV genome references by adding their first 300 bases to the end and mapped the remaining non-human reads to these new HBV reference sequences to identify the genetically closest HBV isolate. In the next two rounds, we mapped the deduplicated reads to 1) the closest HBV isolate and 2) a fine-tuned consensus sequence based on mapping in 1). In the final stage, we cut reads that cross the conventional genome end and combined mappings in the conventional and appended regions. A base was called at the consensus level if allele depth was over x5, with HBV genome coverage calculated by determining the percentage of called consensus bases.

### 2.5 Determining clinically relevant mutations

HBV genotypes, including recombinants, were determined using the online NCBI genotyping tool [36]. Subgenotypes, resistance associated mutations and vaccine escape mutations were called using the online Geno2pheno HBV tool [37].

### 2.6 Software

Graphs were produced using R version 4.2.2 [38], figures were produced with Biorender with a licence to publish [39].

### 2.7 Data availability

Sequences for samples where full length genomes were generated were deposited to the European Nucleotide Archive (ENA) under the study PRJEB79403 for Nanopore data and PRJEB79773 for Illumina.

## 3. Results

### 3.1 Targeted enrichment with probe based capture

#### 3.1.1 Sequencing mock HBV samples enriched with probe based capture using lllumina

We sequenced 10 mock samples (plasmids with genotypes A-J, 5 log_10_ copies/ml) in duplicate, with/without probe-based capture on the Illumina Novaseq X, libraries were diluted to 10pM and pooled prior to sequencing. Without capture, a median of 391 (IQR 153 - 1858) HBV reads per million reads sequenced was generated across 20 mock samples. With capture a median of 563,956 (IQR 397523 - 896635) HBV reads per million reads sequenced was generated (**Supplementary table 1**). Enrichment with probe-based capture led to the generation of full length HBV consensus sequences for all genotypes (**Figure 3A, Supplementary table 1**).

**Figure 3:**
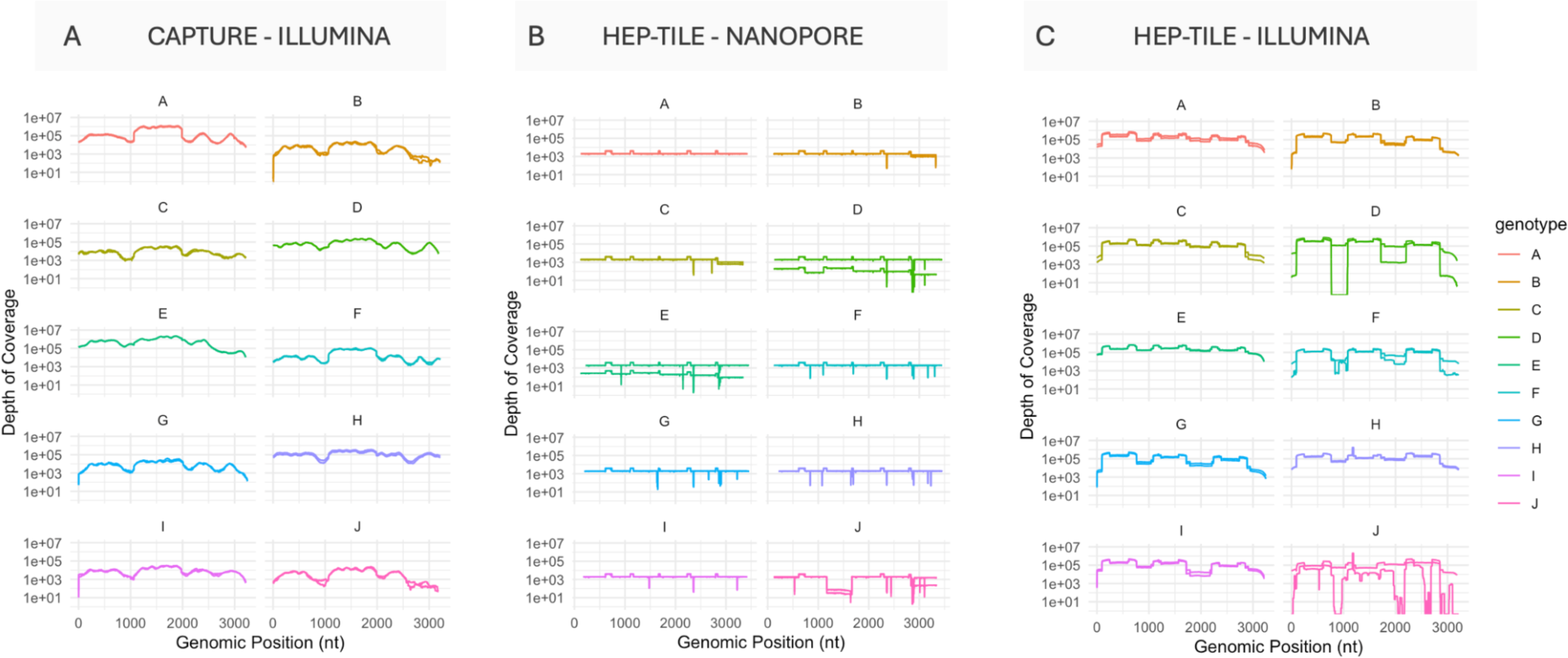
HBV genomes for all HBV genotypes (A-J) in mock clinical samples enriched using HBV probes and primers. (A) Enriched with probe based capture, sequenced on Illumina; (B) Enriched with HEP-TILE, sequenced on Nanopore; (C) Enriched with HEP-TILE, sequenced on Illumina.

#### 3.1.2 Sequencing clinical HBV samples enriched with probe based capture using Illumina

We enriched a subset of 23 clinical samples (HBV VL 4.21 to >7.20 log_10_ IU/ml, as optimisation work established ∼4 log_10_ IU/ml as a conservative estimate for the threshold at which full genomes could be obtained at x5 depth, (**Figure 2**) using probe based capture and sequenced the samples on Illumina Novaseq X. A median of 22,377 (IQR 3267 - 135,184) HBV reads per million reads sequenced was generated. The relationship between HBV VL and percentage reads on target is shown in (**Figure 4A, 4D**). Full length (>98%) genomes at x1 coverage were generated in 18/23 samples, with partial genomes in 5/23. Full length genomes above a minimum x5 read depth (minimum required by bioinformatic pipeline to call base at the consensus level) were generated in 7/23 samples, all of which had VL >5.5 log_10_ IU/ml (∼300,000 IU/ml). Below VL 5.5 log_10_ only partial genomes at x5 depth were generated (**Supplementary table 2**). The negative controls had 0% consensus coverage of the HBV genome.

**Figure 4:**
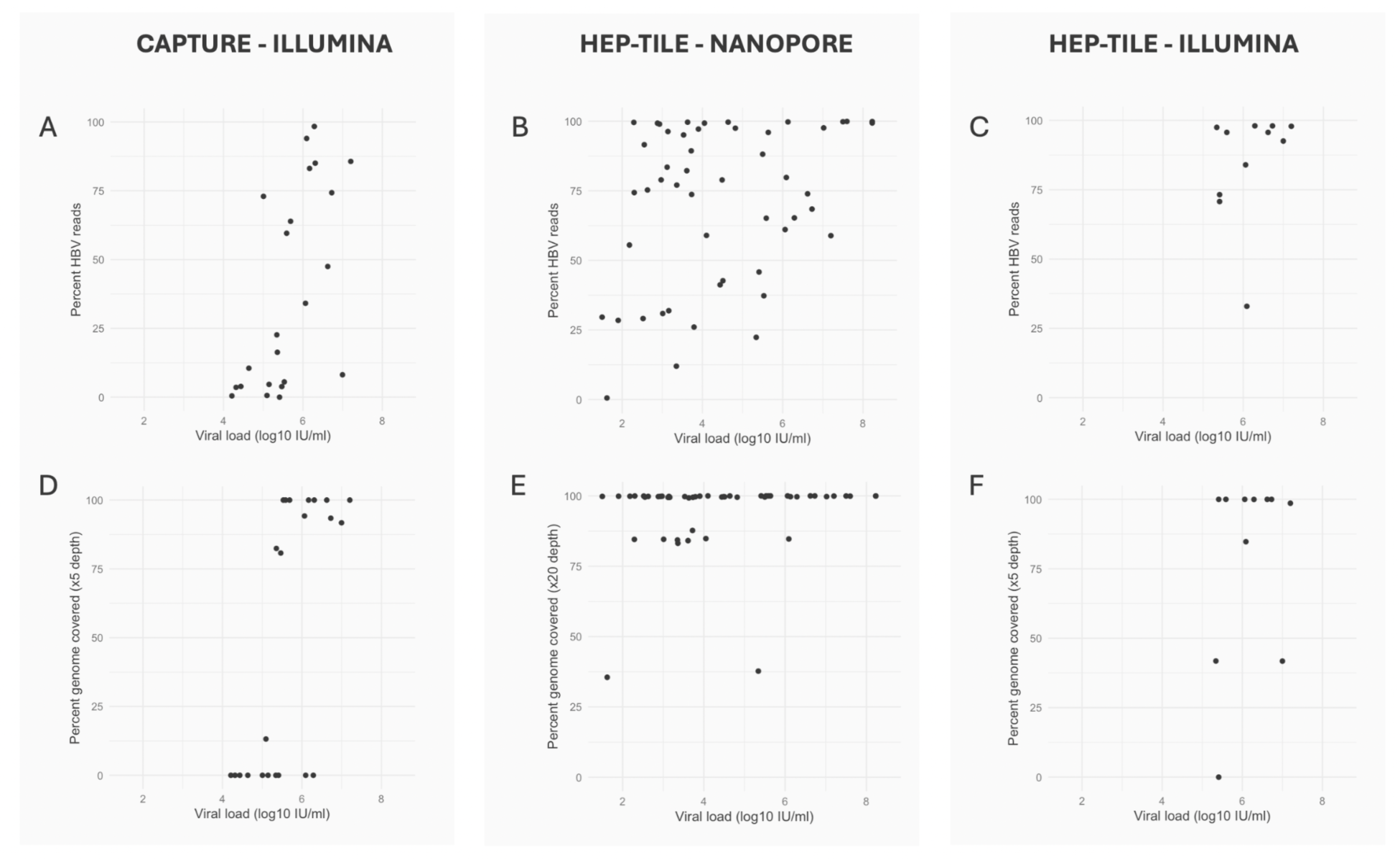
HBV WGS using capture and amplicon based target enrichment. *Top row*: Relationship between viral load and percentage of reads on-target (ie. HBV) for (A) probe-based capture sequenced on Illumina, (B) HEP-TILE sequenced on Nanopore and (C) HEP-TILE sequenced on Illumina. *Bottom row*: Relationship between viral load and percentage of genome with high confidence consensus for (D) probe-based capture sequenced on Illumina (x5 minimum depth), (E) HEP-TILE sequenced on Nanopore (x20 minimum depth) and (F) HEP-TILE sequenced on Illumina (x5 minimum depth). Panels E and F show an outlier with viral load 5.41 log10 IU/ml, with disproportionately low percentage genome coverage. Repeat VL was 3 log10 IU/ml.

### 3.2 Targeted enrichment with HEP-TILE tiled amplicon PCR

#### 3.2.1 Sequencing mock HBV samples enriched with HEP-TILE using Nanopore and Illumina

We sequenced the same 10 mock samples in duplicate, enriching using the HEP-TILE tiled amplicon scheme and sequenced on Nanopore MinION and Illumina Novaseq X. With HEP-TILE Nanopore a median of 997,016 (IQR 994,669 - 999,427) HBV reads per million reads sequenced was generated. With HEP-TILE Illumina a median of 977,441 (IQR 963,459 - 982,631) HBV reads per million reads sequenced was generated (**Supplementary table 1**). Enrichment with HEP-TILE led to the generation of full-length HBV sequences across genotypes on both platforms (**Figure 3B, 3C**).

#### 3.2.2 Sequencing clinical HBV samples enriched with HEP-TILE using Nanopore

We enriched a subset of 50 clinical samples (HBV VL 1.50 to >8.23 log_10_ IU/ml, **Figure 2**) using the HEP-TILE protocol and sequenced on the Nanopore MinION. A median of 779,909 (IQR 590,906 - 676,536) HBV reads per million reads sequenced was generated. The relationship between HBV VL and percentage reads on target is presented (**Figure 4B, 4E**).

Full genome coverage (x1 depth) was generated in 40/50 samples. Full-length HBV consensus genomes at x20 depth (the minimum required to accurately call a mutation), were generated in 40/50 samples, down to the lowest VL tested (1.5 log_10_ IU/ml). Partial genomes were obtained for the remaining 10 samples. These low coverage samples tended to have lower VL (**Figure 4**), although this did not reach statistical significance (p = 0.16, Mann Witney U test). We did not observe any relationship between HBV genotype and inability to produce whole genomes (**Supplementary figure 2**). The negative controls had 0% consensus coverage of the HBV genome.

#### 3.2.3 Sequencing clinical HBV samples enriched with HEP-TILE using Illumina

We selected a subset of 10 samples (based on sample availability) for HEP-TILE amplification, followed by fragmentation and sequencing on Illumina (Novaseq X 150PE partial lane) (**Figure 2**). A median of 956,800 (IQR 759,122 - 977,744) HBV reads per million reads sequenced was generated. The relationship between HBV VL and percentage reads on target is shown in (**Figure 4C, 4F**). Full genome coverage (x1 depth) was generated in 7/10 samples. Full-length consensus HBV genomes (minimum x5 read depth) were generated in 7/10 samples, partial genomes were obtained for 2/10 samples and the pipeline failed to generate a consensus from 1 sample due to low read coverage and anomalous mapping mismatches. The negative controls had 0% consensus coverage of the HBV genome.

### 3.3 Methods comparison

#### 3.3.1 Accuracy of consensus genomes

We sequenced all 10 mock samples using all three workflows, with each sample processed in duplicate (capture Illumina, HEP-TILE Illumina, HEP-TILE Nanopore). Comparison of consensus sequences generated from mock samples to the known plasmid sequence, demonstrated 100% accuracy of consensus genomes produced with capture-Illumina and HEP-TILE Nanopore and 99.98% with HEP-TILE Illumina (**Table 1**). Four samples sequenced with HEP-TILE Illumina had mismatches or bases designated ‘N’, however the median number of mismatches and N’s was 0. There were no indels for any of the methods.

**Table 1:**
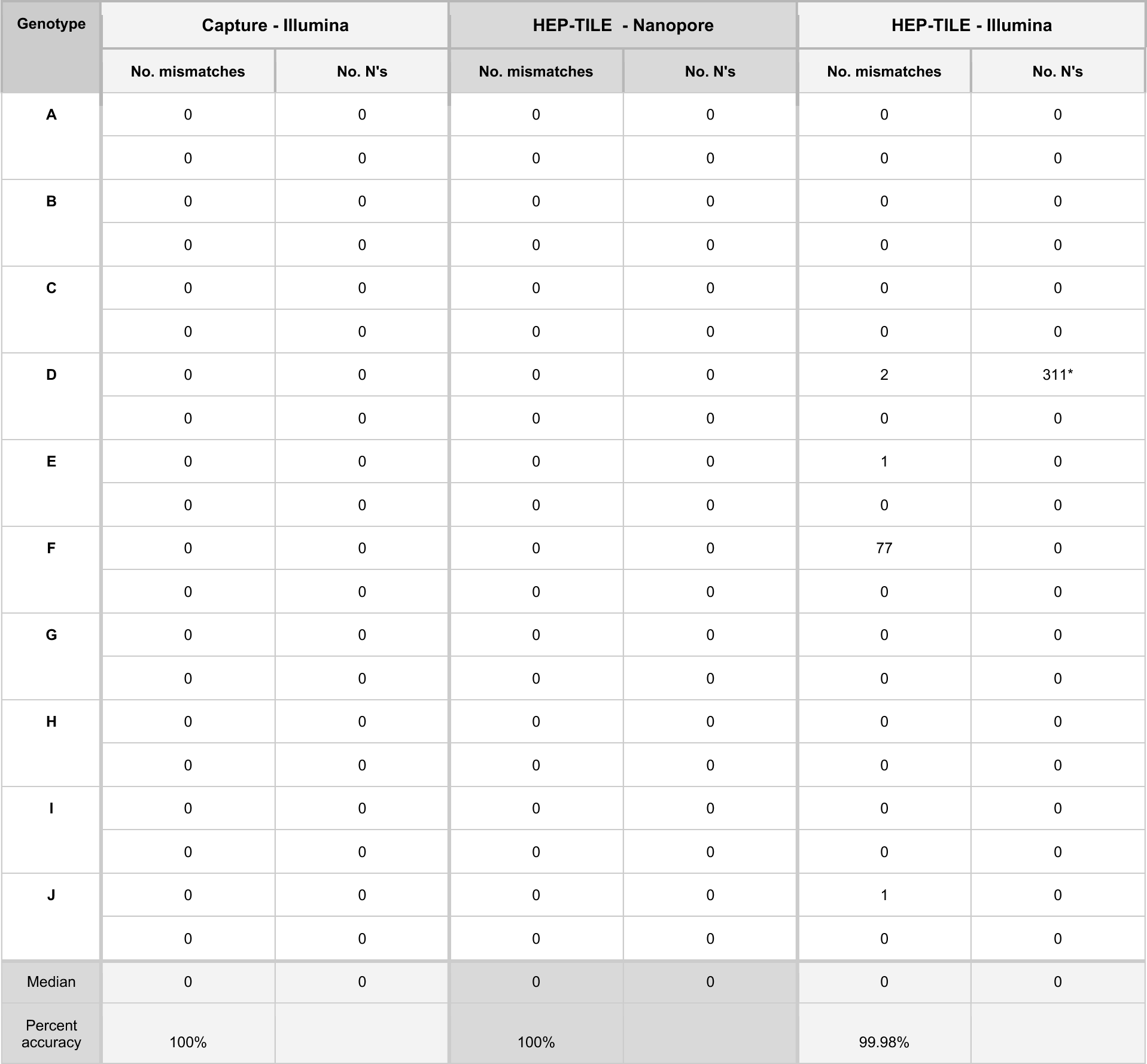
Comparison of sequencing errors when sequencing mock samples using three workflows. Each mock sample was sequenced in duplicate, one replicate per row. * 311 base dropout of a continuous regio

#### 3.3.2 Comparison of capture vs. HEP-TILE ability to generate whole genomes from high viral load clinical samples

We sequenced a subset of 10 clinical samples (> 5 log_10_ IU/ml) using all three workflows so that we could compare the ability of capture vs. HEP-TILE to generate HBV WGS at high VL. At x1 depth, capture-Illumina and HEP-TILE Nanopore generated full genomes in 8/10 samples, with HEP-TILE Illumina in 7/10 samples. At depths required to confidently call consensus (x5 for Illumina, x20 for Nanopore), full genomes were generated in 4/10 samples for capture-Illumina, 8/10 for HEP-TILE Nanopore and 7/10 for HEP-TILE Illumina (**Supplementary table 2**).

### 3.4. Determining genotypes from HBV WGS data

We identified samples of genotype A-E, in addition to genotype B/C and D/E recombinants (see methods) (**Supplementary table 2**). We were able to call polymorphisms at sites of previously reported resistance associated mutations (RAMs) and vaccine escape mutations (VEMs) [37] (details in **Supplementary text**).

## 4. Discussion

### 4.1 Summary of outputs

We have developed two pan-genotypic viral enrichment methods for HBV WGS: HEP-TILE and probe based capture. Both techniques effectively enrich HBV genomes from all known genotypes (A-J), as evidenced by application to mock samples, and increase the percentage of sequencing reads mapping to the HBV genome, therefore reducing the amount of sequence data needed to obtain full genome coverage. We used HBV WGS to call HBV genotypes, including recombinants, which may be missed with partial-genome sequencing. HEP-TILE capitalises on the rapid global expansion of technical expertise and infrastructure for amplicon based WGS that occurred during the SARS-CoV-2 pandemic. Having a toolbox of diverse sequencing methods allows the most appropriate method to be selected according to local infrastructure, cost and throughput requirements (**Table 2**).

**Table 2:** Summary of three HBV WGS pipelines, comparing time, cost and outputs. All costs are based on cost of reagents in the UK in 2024 (GBP) and do not include staff or equipment costs. *based on sequencing of mock samples with known sequence ** reagent costs for enrichment and library prep reagents calculated in GBP for first 1000 samples. Cost excludes extraction and QC (Qubit/Bioanalyser reagents). *** NB price extremely variable - depends on platform used and whether performing in-house or outsourcing commercially. Excludes hardware (i.e. Laptop/MinION for Nanopore, and Novaseq/Miseq for Illumina)

|  | Capture - Illumina | HEP-TILE - Nanopore | HEP-TILE - Illumina |
| --- | --- | --- | --- |
| Time required for enrichment and library prep | 1.5 days (capture overnight) | 1.5 days (PCR overnight) | 1.5 days (PCR overnight) |
| Sensitivity (HBV DNA VL) for generating HBV WGS | > 5.5 log<sub>10</sub> IU/ml | > 1.5 log<sub>10</sub> IU/ml | > 1.5 log<sub>10</sub> IU/ml |
| Consensus sequence accuracy* | 100% | 100% | 99.98% |
| Cost per sample** (enrichment & library prep reagents) | £29 | £17 | £25 |
| Cost per sample*** (sequencing) | £25<br>E.g.multiplexed on partial lane on Novaseq X, 1 million reads per sample | £5.60<br>E.g. 96 samples multiplexed per flow cell, bulk purchasing 12 flow cells, 100,000 reads per sample | £2.50<br>E.g. multiplexed on partial lane on Novaseq X, 100,000 reads per sample |
| Throughput | Platform dependent e.g. Miseq vs. Novaseq X | Platform dependent e.g. Minion vs. Promethion | Platform dependent e.g. Miseq vs. Novaseq X |
| Portability | No | Yes with MinION | No |

### 4.2 Comparison between methods

HEP-TILE offers the greatest flexibility to sequence across a range of HBV VL, able to sequence full genomes down to 1.50 log_10_ IU/ml (∼30 IU/ml, lowest tested here), using standard extraction from 200µL - 400µL plasma. The primers are designed to be pan-genotypic (A-J), unlike other current schemes designed for genotypes A-E [40,41]. The ∼600-715bp amplicons used in HEP-TILE improve sensitivity compared to single amplicon schemes [42], enhancing sequencing success from low VL or degraded samples. While HEP-TILE amplicons can be sequenced on either Nanopore or Illumina, the use of the ONT native barcoding kit described here (ligation based) rather than ONT rapid barcoding kit or NEB-Ultra-II-FS kit (both requiring fragmentation) enables the sequencing of whole amplicons. Although shorter amplicon schemes (300-600bp) can facilitate transfer to Illumina platforms without amplicon fragmentation, HBV diversity and genome structure prohibit the generation of a shorter pan-genotypic scheme (**Supplementary figure 1**). Generally, shorter amplicons improve sensitivity and platform transferability, however longer 1-2 amplicon schemes or schemes generating concatenated full genomes are useful for haplotype reconstruction [4,40]. The use of two primer pools (instead of individual reactions for each amplicon) simplifies the PCR step and eliminates the need for re-balancing of individual primer pools post-PCR required by other methods [43].

We report percent genome coverage at x1 depth in addition to the percentage of the genome with consensus calls to allow comparison with other published methods presenting coverage at x1 depth. We use a higher threshold of x20 depth to call bases at a consensus level for Nanopore, and x5 for Illumina to improve certainty of the consensus base call (higher threshold for Nanopore used due to the higher error rate of Nanopore sequencing technology).

**Table 2** compares the costs of each workflow, estimated per sample based on an initial batch of 1000 samples. Cost per sample decreases with multiplexing (e.g. 96 samples for Nanopore, more possible for Illumina). Due to the initial cost of purchasing capture probes (∼£6300 for this panel), sequencing smaller sample sets with capture becomes prohibitively expensive. Costs for Illumina sequencing vary significantly depending on whether the option of purchasing a partial lane on a high throughput platform is available.

### 4.3 Real world application of methods

Although full genome HBV sequencing coverage should be achievable for the majority of samples using HEP-TILE, if samples at extreme ends of the VL spectrum are pooled on the same run, uneven read distribution is expected between samples, with loss of coverage for the lowest VL samples. Therefore batching samples by VL and normalising the mass of PCR product being taken forwards into library preparation is recommended. Viral enrichment by PCR is highly sensitive to amplicon contamination from previous experiments therefore caution should be taken to keep work areas, equipment and reagents free of contamination, and multiple negative controls should be included. To avoid *in-silico* ‘contamination’ double barcoding is required when demultiplexing to eliminate chimeric reads which may otherwise lead to mis-assignment of reads to incorrect barcodes [44]. These wet lab and bioinformatic principles are also applicable to sequencing other targets.

Although our capture approach can be used to improve the efficiency of viral sequencing at VL >∼5.50 log_10_ IU/ml, its efficacy for producing full-length consensus genomes diminishes at lower VLs, as observed elsewhere [45]. Additional/alternative host depletion and high volume (e.g. 5ml [46]) extraction is likely to be required to reliably obtain full consensus genomes using this method in samples with VL <5.5 log_10_ IU/ml.

### 4.4 Caveats and limitations

The results presented here are from an iterative set of experiments and protocol development, rather than a pre-planned head-to-head comparison. This limits the direct comparability of workflows due to insufficient sample volumes of the original South Africa sample set. This meant we were unable to sequence all samples using all three enrichment approaches. Furthermore, the extraction method changed over time. Nevertheless, these data showcase the capabilities of each workflow.

Correlation between VL and percentage genome coverage was more variable than anticipated. Factors such as storage duration, shipping conditions, and repeated freeze-thaw cycles may all influence sample quality, although DNA is generally robust. Poor sample quality or inaccurate VL reporting (different sample/timepoint tested or lab/reporting error) may explain instances where samples that were initially reported to have high VL yielded lower coverage than expected (e.g. sample 1116 for which VL was reported as 5.41 log_10_ IU/ml but on repeat on our sample was 3 log_10_ IU/ml).

The probes and primers were designed to be pan-genotypic, however only take into account the diversity of sequences on HBVdb, which contains relatively few genotype F-H sequences, and under-represents certain global populations [ref Delphin?]. A strength of our study is the use of mock samples, which allowed us to assess performance across all genotypes, including rarer genotypes F-J that were unlikely to be present in our cohorts, however might still be present in other cohorts globally. Analysis of rarer genotypes is often missing in reports of sequencing schemes, however will become increasingly important as more HBV WGS is performed from diverse global regions, and known HBV sequence diversity expands. While we report these methods as pan-genotypic using mock samples generated from plasmids containing HBV genotypes A-J, it is important to note that we only tested one variant per genotype. Some genotypes have multiple subtypes; future studies could test enrichment methods with a wider range of subgenotypes.

Sequencing low VL samples has inherent stochasticity, depending on the presence of viral DNA in a sample or extract, and assay chemistry. Future work will test HEP-TILE performance at viral loads <1.5 log_10_ IU/ml, using high volume extraction to increase the chance of sampling viral DNA (e.g. extraction from 5ml serum/plasma [46]), increasing the number of PCR cycles from 35 to 40 and pooling products from PCRs performed in duplicate or triplicate. Future work could compare the ability of capture or HEP-TILE to accurately quantify low abundance minority variants, requiring a panel of control material at varying VLs with known variant percentages, sequenced with multiple technical replicates. Finally, enrichment methods could be trialled on different sample types, for example liver biopsies where high host background is a challenge.

### 4.5 Conclusion

Collectively, these protocols will facilitate the wider generation of HBV sequence data. This data will lead to a more accurate and representative global picture of HBV molecular epidemiology, cast light on persistence and pathogenesis, and enhance understanding of treatment responses and clinical outcomes. These aspirations align with high profile international goals for elimination of HBV as a public threat.

## Data Availability

All data produced in the present work are contained or linked in the manuscript

## Author contributions

Conceptualization: SFL, JQ, MAA, PCM

Methodology: SFL, CK, DJ, HC, AT

Software: CK, HC, SW, JQ, MAA

Formal Analysis: CK, HC, SFL

Investigation: SFL, CK, DJ, HC, GA, EW, MD, AT, SW, YW, G M-C, MK, KO, JC

Resources: BK, TGM, CdL, JM, MVS, DG, RN, EB, PCM

Data curation: CK, HC, JQ, MAA

Writing (original draft): SL, CK

Writing (review and editing): all authors

Visualisation: SL, CK

Supervision: JQ, MAA, PCM

Funding acquisition: SFL, JQ, MAA, PCM

## Data availability statement

Data for this study is available within the manuscript, supplementary table and online links. Sequences for samples where full length genomes were generated were deposited to the European Nucleotide Archive (ENA) under the study PRJEB79403 for Nanopore data and PRJEB79773 for Illumina.

## Supplementary material

**Supplementary figure 1:**
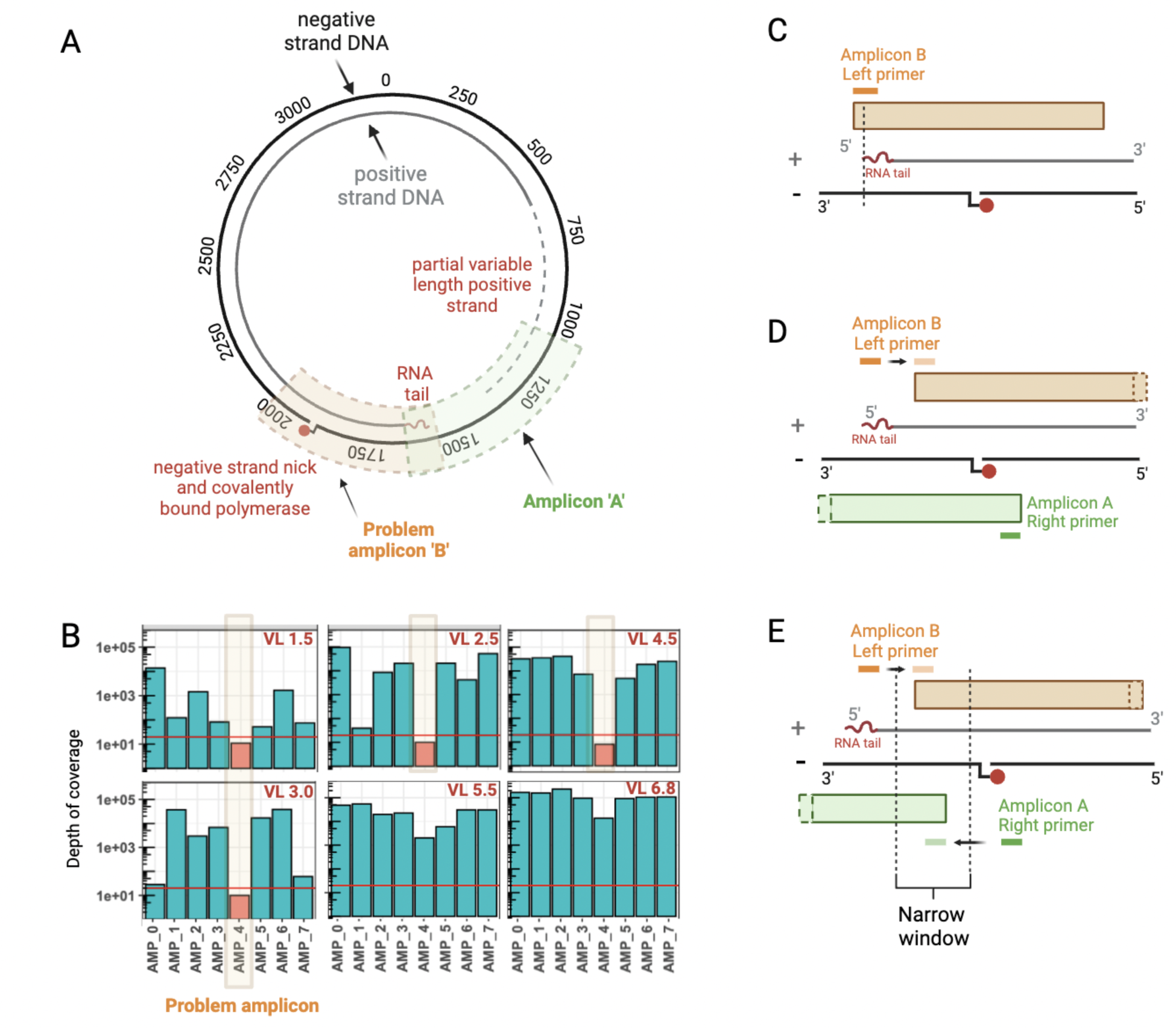
Modifications to primer positions to take into account HBV genome structure. (A) Schematic of HBV relaxed circular DNA (rc-DNA), the HBV partially dsDNA genome has 5’ modifications to both the positive DNA strand (RNA tail) and negative DNA strand (covalently bound polymerase). The negative strand is complete but not fully circular due to the presence of a ‘nick’ and covalently bound polymerase. The partial positive strand is variable in length. (B) Depth of coverage for amplicon scheme hbv/500/v1.1.0 - illustrating drop-out of the ‘problem amplicon’ (initially Amplicon 4) in samples below a VL of 5 log10 IU/ml (VL shown in red). (C) Amplicon B drops out in low-medium viral load samples, the hypothesis is that neither the positive nor negative strands of the HBV relaxed circular genome are complete in that region. The left hand primer only partially overlaps the 5’ tail of the positive strand, the negative strand contains a nick. (D) Attempt to solve issue by moving the amplicon B primer upstream, however this has the knock on effect of the Amplion A right hand primer moving upstream, which now means amplicon A falls across a non-continuous DNA region. (E) Solution requires the left hand primer for amplicon B and right hand primer for amplicon A to be placed in a narrow 200bp window, to ensure Amplicon B can be generated from the positive DNA strand, and amplicon A from the negative DNA strand. Created in BioRender. Lumley, S. (2024) BioRender.com/l10u171

**Supplementary figure 2:**
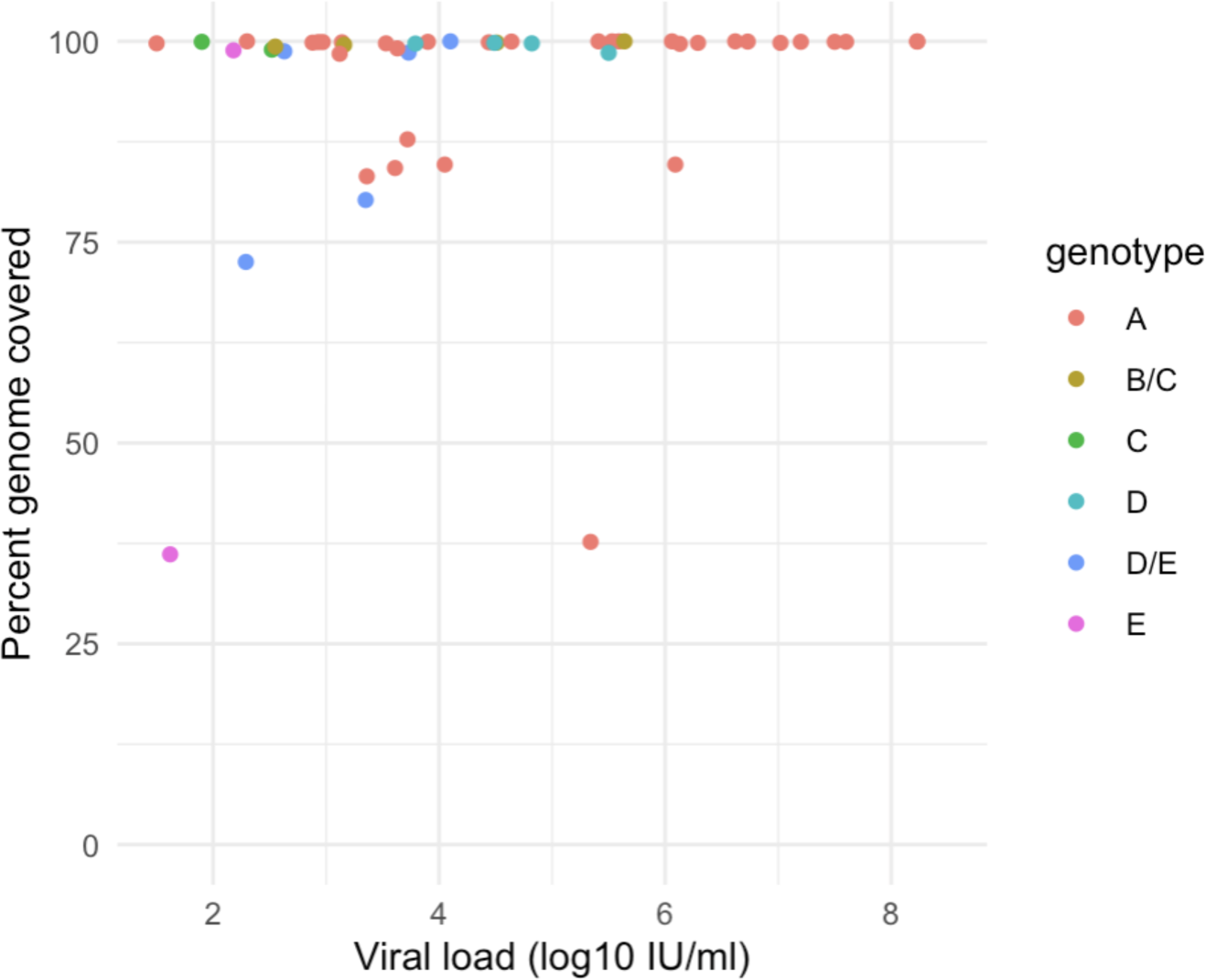
Relationship between HBV genotype and performance of HEP-TILE Nanopore workflow. Percentage genome coverage shown at x20 depth.

**Supplementary table 1:**
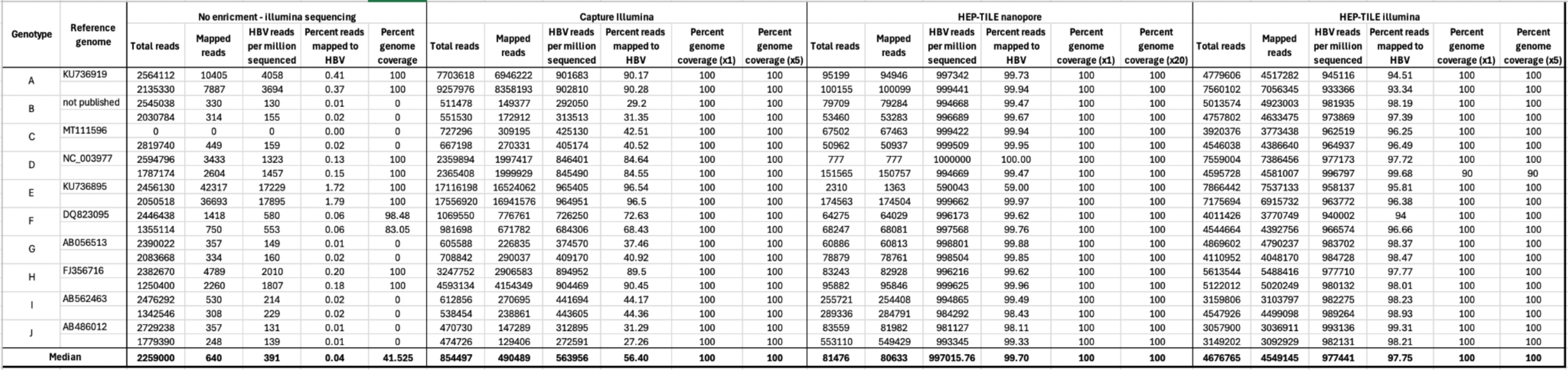
Sequencing metrics for mock HBV samples for capture-Illumina, HEP-TILE Nanopore and HEP-TILE Illumina workflows vs. no enrichment.

**Supplementary table 2:**
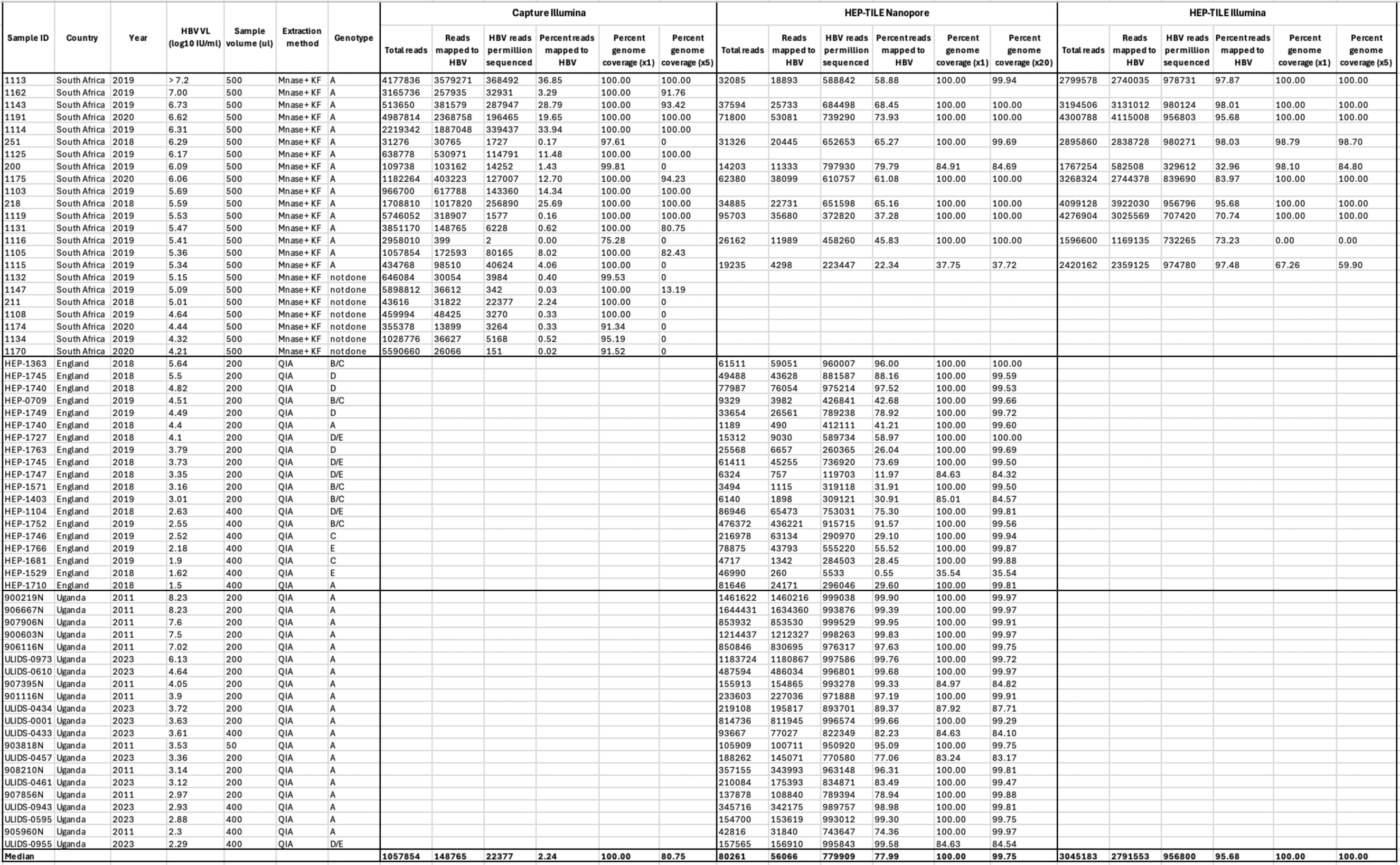
Metadata and sequencing outputs for clinical HBV samples sequenced with capture-Illumina, HEP-TILE Nanopore and HEP-TILE Illumina workflows. Sequencing metrics for sequencing of HBV-positive clinical samples. Key for extraction methods (see methods section for full details): “Mnase + KF” = host depletion with micrococcal nuclease followed by extraction on Kingfisher apex, “QIA” = QiaAMP MinElute virus spin kit. Viral load conversion example: 2log10 IU/ml = 100 IU/ml, 3log10 IU/ml = 1000 IU/ml.

**Supplementary table 3.** Number of sequences downloaded from publicly available sequence database HBVdb. [20]

| Genotype | Capture<br>(2019 HBVdb download) |  | HEP-TILE<br>(2024 HBVdb download) |  |
| --- | --- | --- | --- | --- |
|  | Genomes in<br>HBVdb | Genomes used | Genomes in<br>HBVdb | Downsampled<br>Genomes |
| A | 506 | 506 | 1066 | 559 |
| B | 1218 | 1218 | 2018 | 1143 |
| C | 1447 | 1447 | 3095 | 1764 |
| D | 823 | 823 | 1523 | 920 |
| E | 254 | 254 | 397 | 254 |
| F | 197 | 197 | 304 | 156 |
| G | 28 | 28 | 50 | 18 |
| H | 26 | 26 | 28 | 17 |

**Supplementary table 4.**
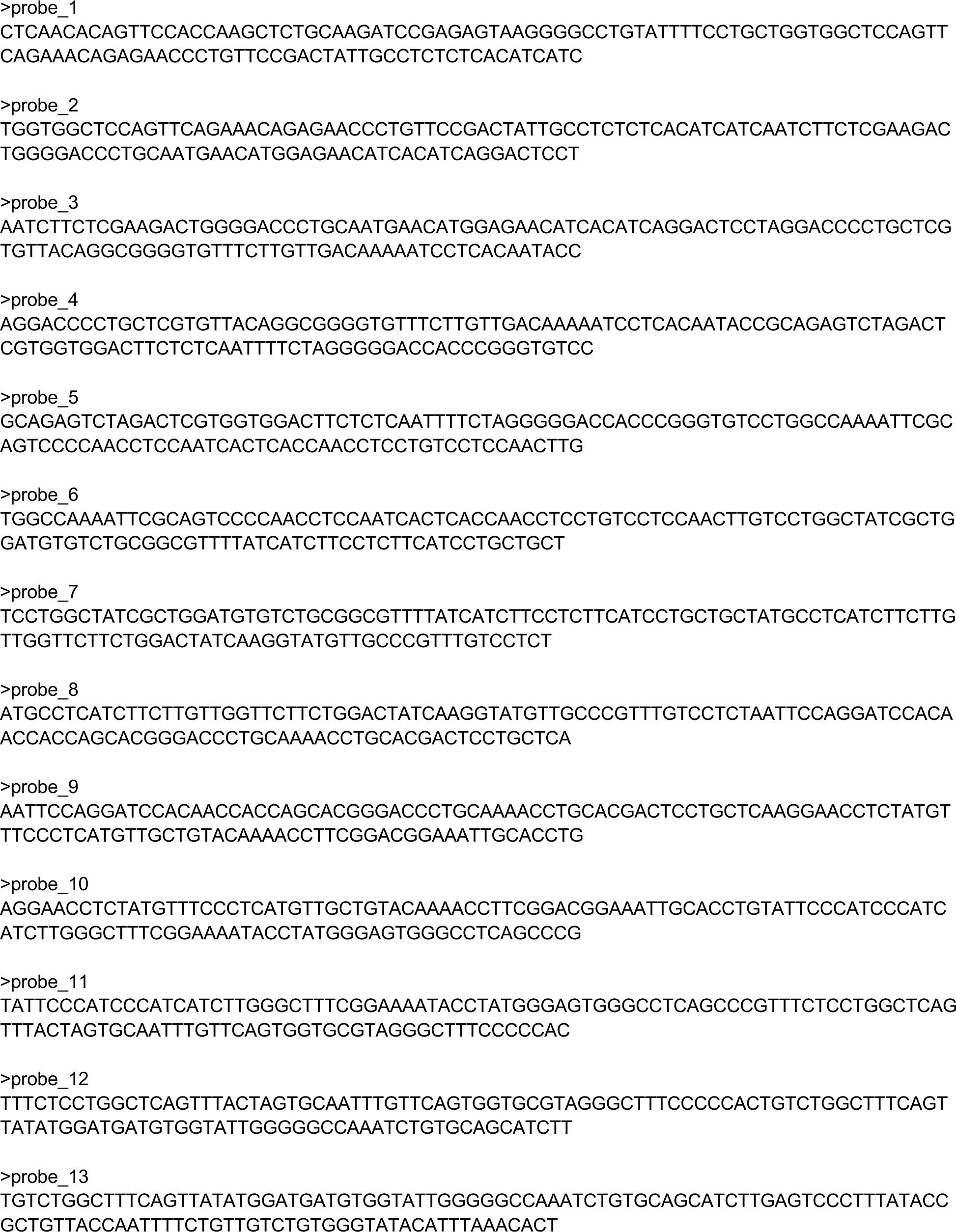
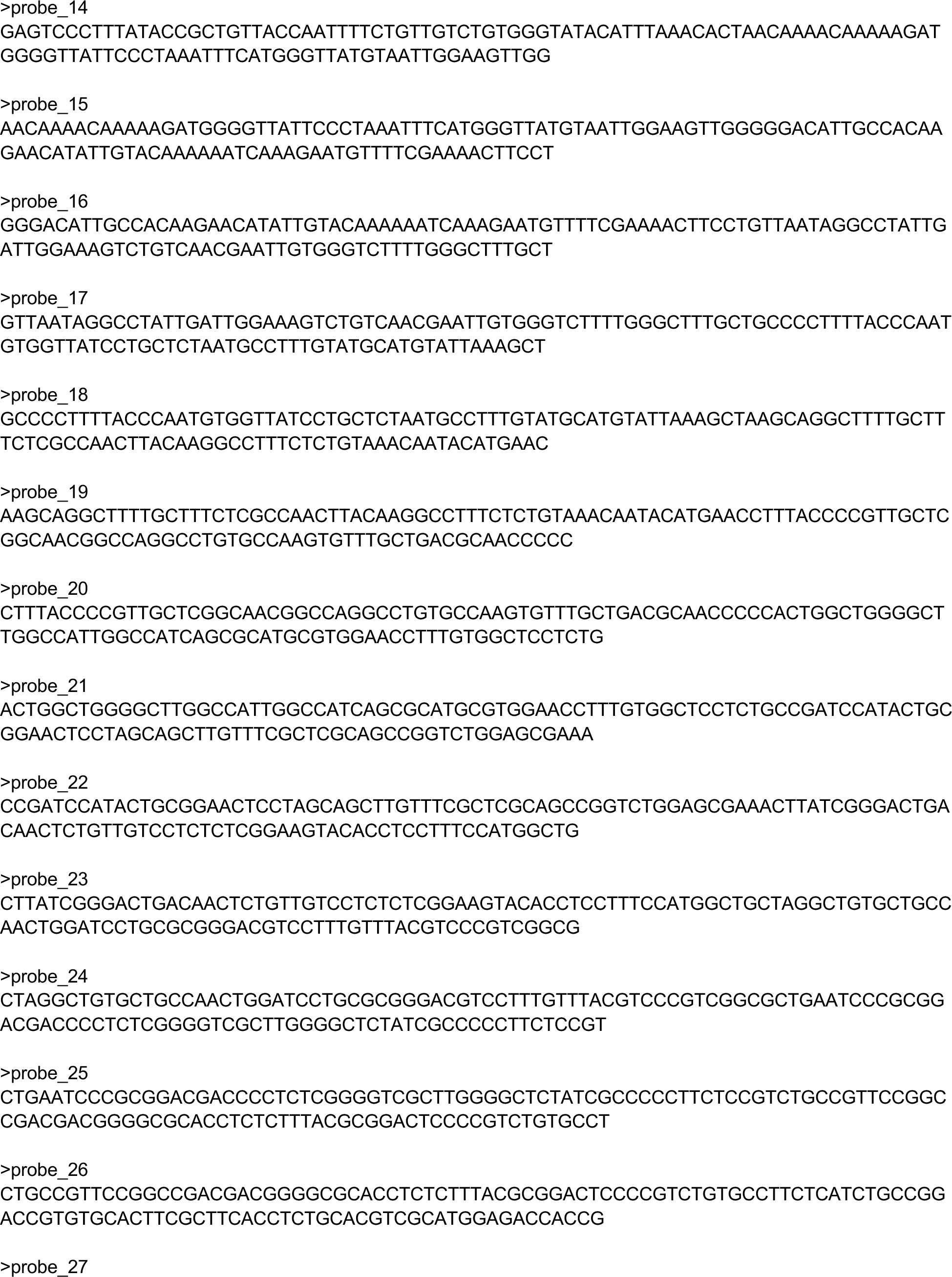
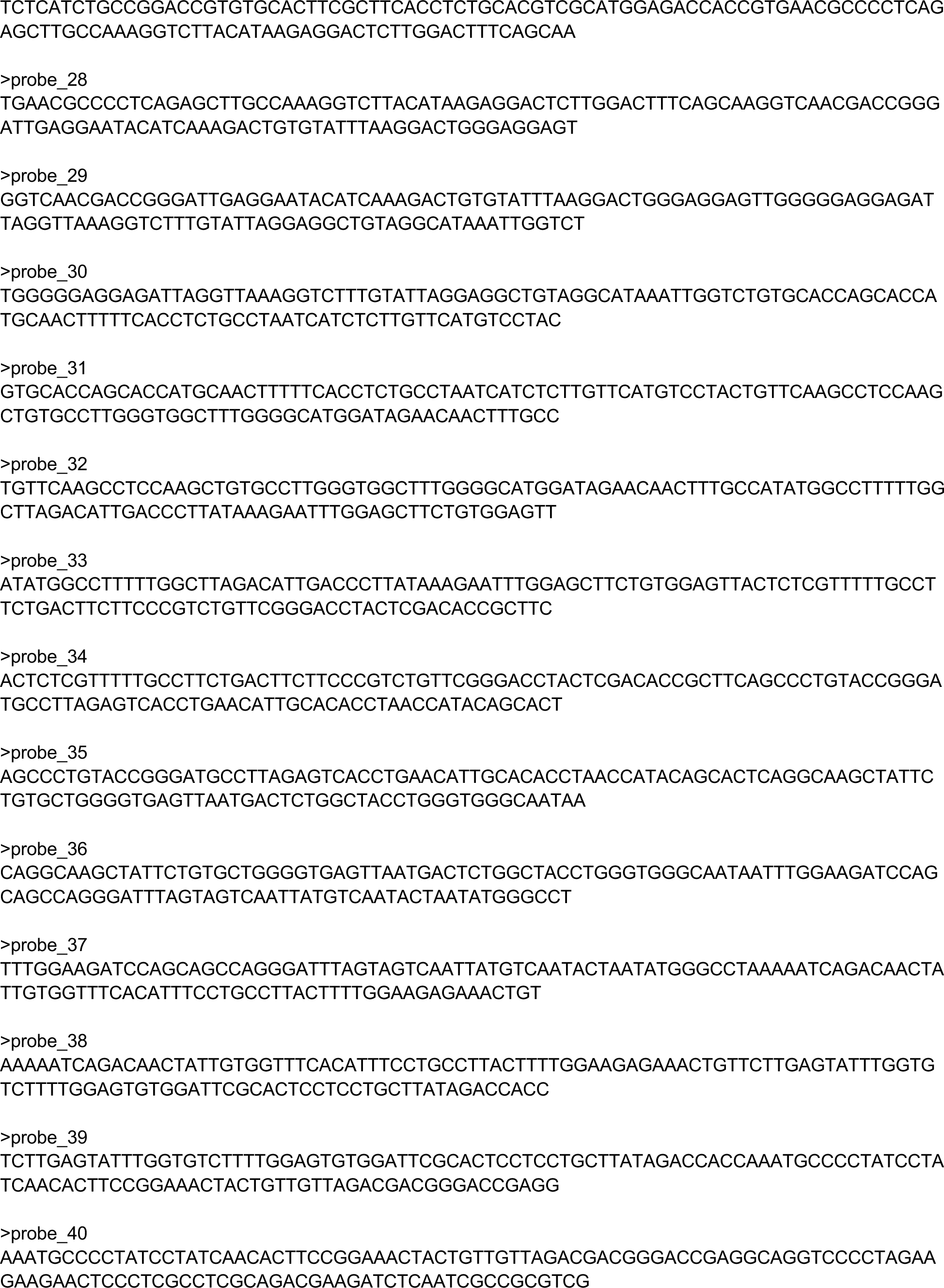
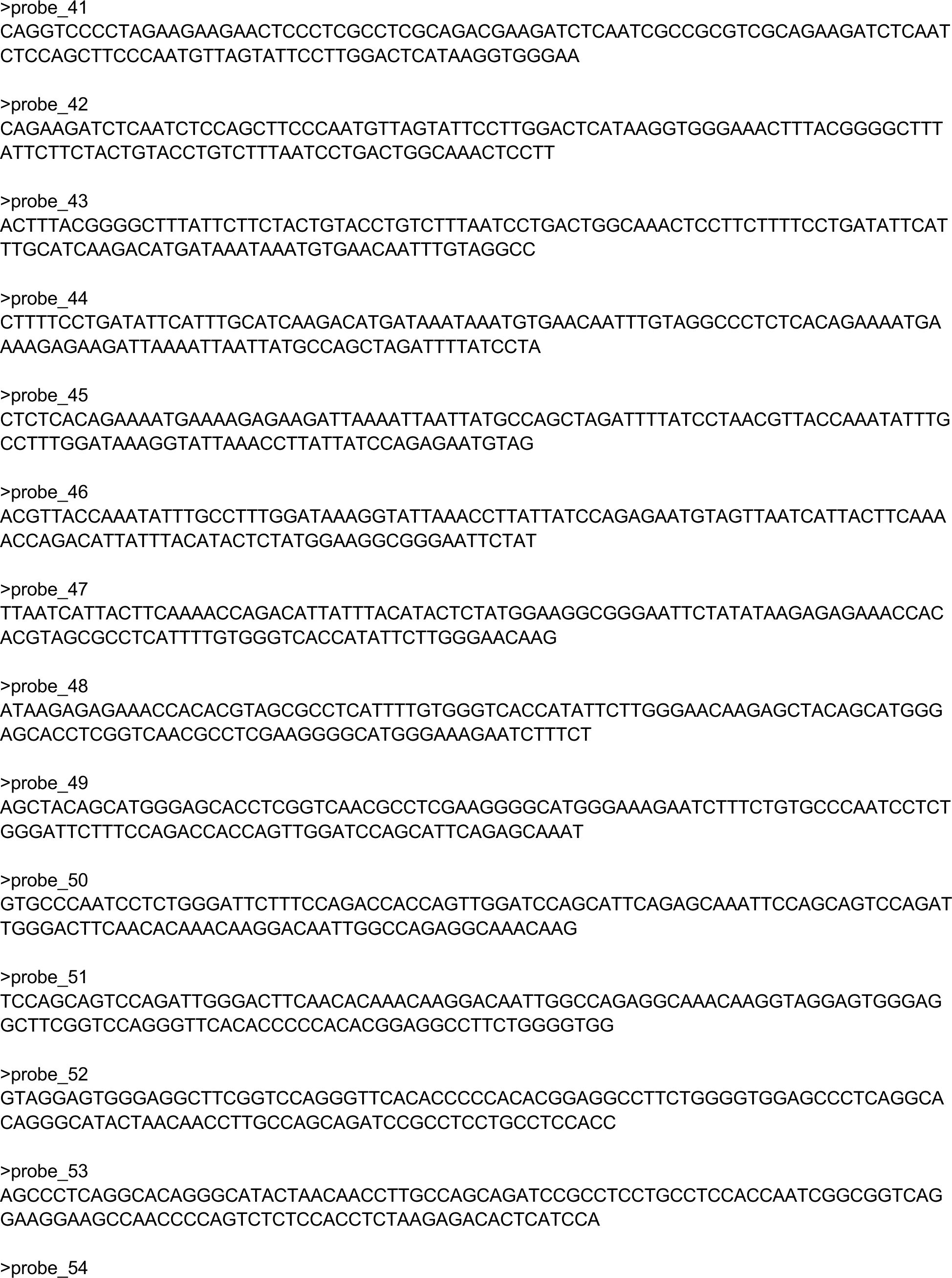
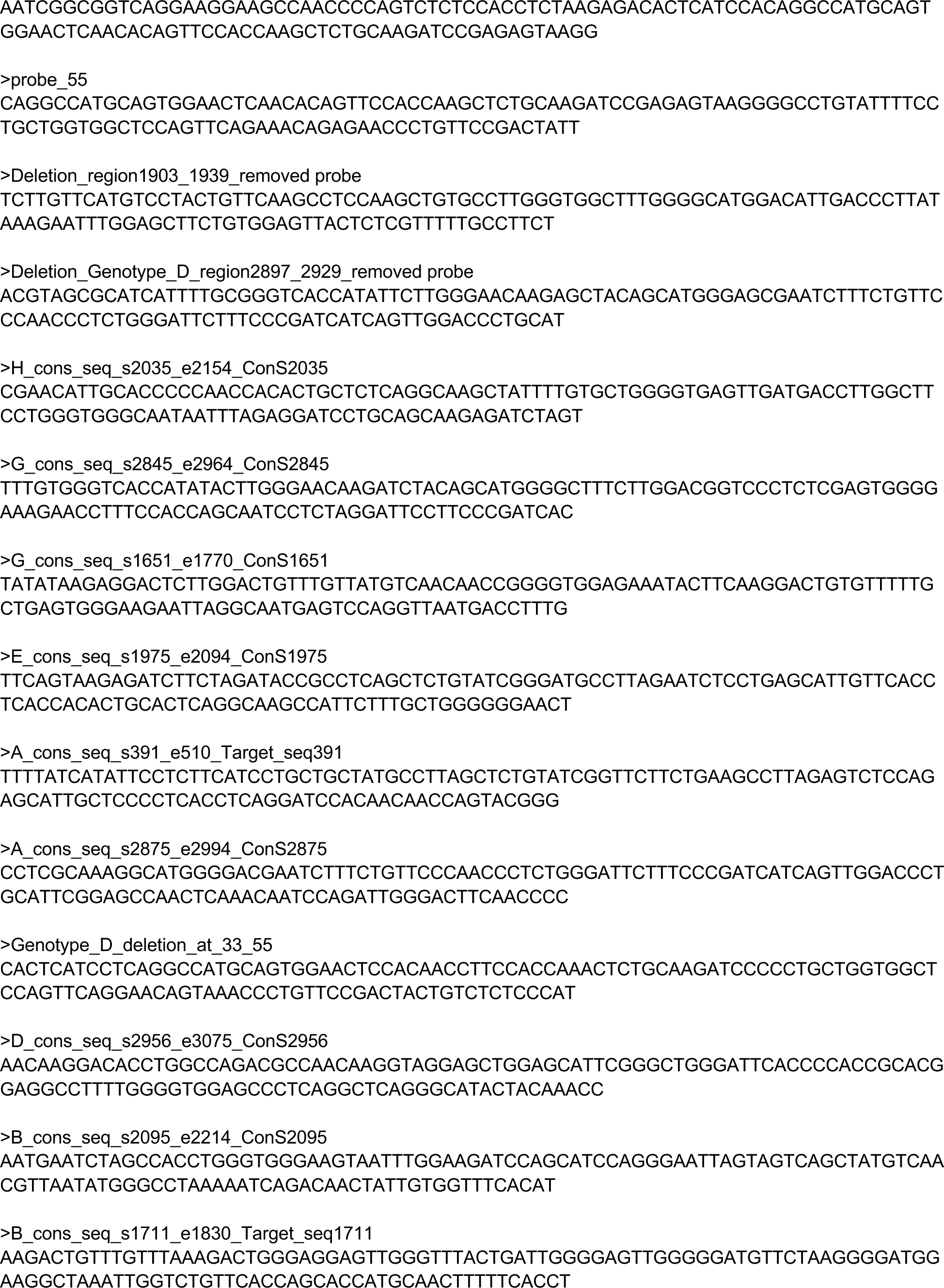
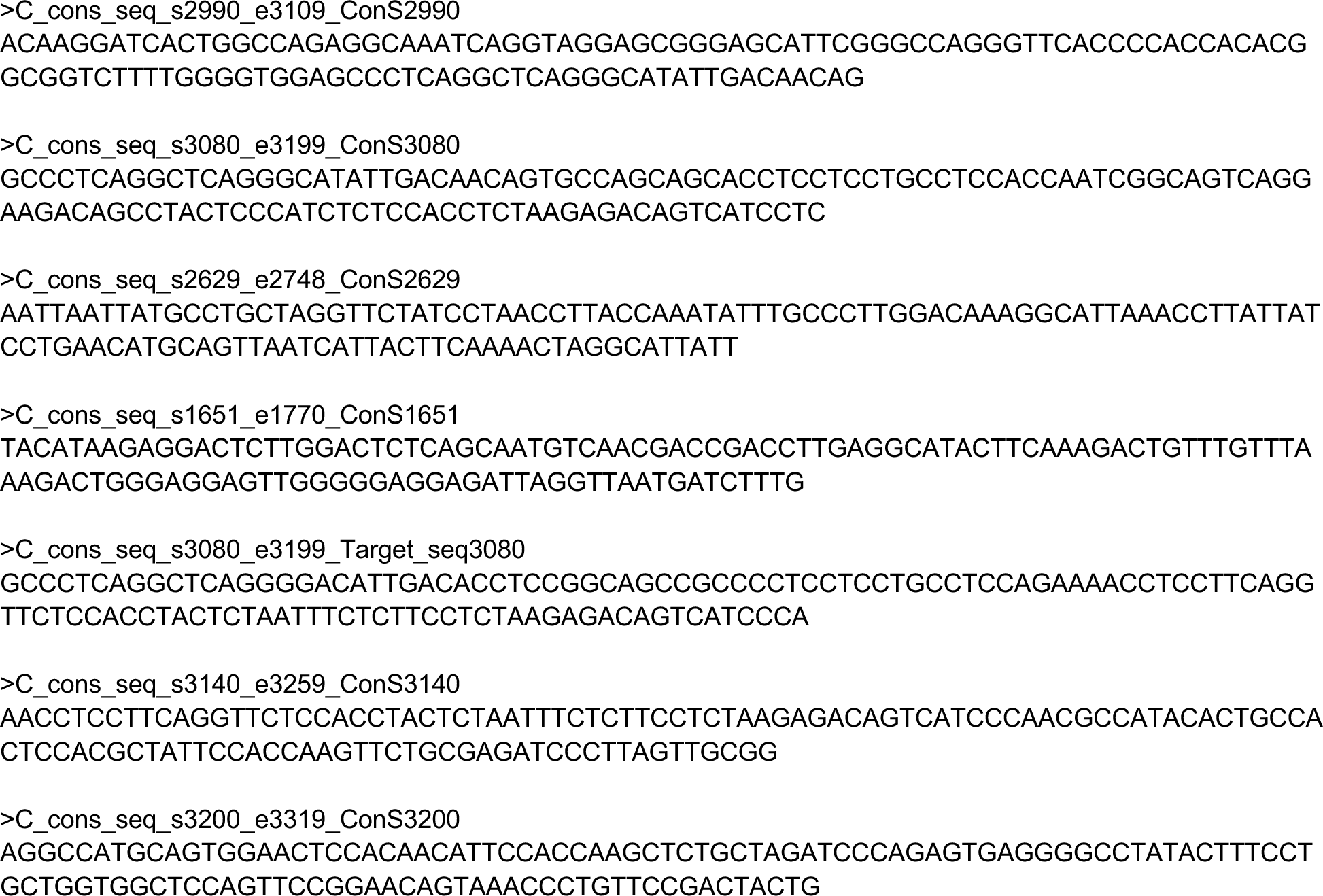
HBV probe sequences.

## Supplementary text

### Determining HBV drug resistance and immune escape from sequences using HBV WGS

We were able to call polymorphisms at sites of previously reported resistance associated mutations (RAMs) and vaccine escape mutations (VEMs) as defined by geno2pheno[37]. Polymorphisms were detected in RT and HBsAg regions. The polymorphism rt169M was identified in viral sequences from two individuals (906116N/907395N) and 202R in one (HEP-1763). The rt169 and rt202 are sites associated with HBV drug resistance however the mutations 169M and 202R have unknown significance. In the HBsAg region, a number of mutations associated with HBsAg escape from vaccines and detection were identified (144E, 100C, 128V, 133I and 134V, each in one individual). All samples with polymorphisms had only been sequenced with a single method, HEP-TILE Nanopore.

